# Patient-specific hemodynamic markers co-localise with regions of neointimal hyperplasia in an arteriovenous graft

**DOI:** 10.1101/2024.12.05.24318387

**Authors:** Federica Ninno, Catriona Stokes, Edouard Aboian, Alan Dardik, David Strosberg, Stavroula Balabani, Vanessa Díaz-Zuccarini

## Abstract

**Purpose:** Most computational fluid dynamics (CFD) studies on arteriovenous grafts (AVGs) adopt idealised geometries and simplified boundary conditions (BCs), potentially resulting in misleading conclusions when attempting to predict neointimal hyperplasia (NIH) development. Moreover, they often analyse a limited range of hemodynamic indices, lack validation, and fail to link the graft-altered hemodynamics with follow-up data. This study develops a novel patient-specific CFD workflow for AVGs using pathophysiological BCs. It validates the CFD results with patient medical data and assesses the co-localisation between CFD results and NIH regions at follow-up.

**Methods:** Contrast-enhanced computed tomography angiography images were used to segment the patient’s AVG geometry. A uniform Doppler ultrasound (DUS)-derived velocity profile was imposed at the inlet, and three-element Windkessel models were applied at the arterial outlets of the domain. Transient, rigid-wall simulations were performed using the k-ω SST turbulence model. The CFD-derived flow waveform was compared with the patient’s DUS image to ensure validation. Turbulent kinetic energy (TKE), helicity and near-wall hemodynamic descriptors were calculated and linked with regions presenting NIH from a 4-month follow-up fistulogram.

**Results:** In the analysed patient, areas presenting high TKE and balanced helical flow structures at baseline exhibit NIH growth at follow-up. Transverse wall shear stress index is a stronger predictor of NIH than other commonly analysed near-wall hemodynamic indices, since luminal areas subjected to high values greatly co-localise with observed areas of remodelling.

**Conclusion:** This patient-specific computational workflow for AVGs could be applied to a larger cohort to unravel the link between altered hemodynamics and NIH progression in vascular access.

## 1. Introduction

Chronic kidney disease (CKD) is a progressive pathology affecting more than 800 million people worldwide [1]. In end-stage renal disease, patients rely on hemodialysis as the primary form of renal replacement therapy to survive [2]. Hemodialysis involves supplementing the kidney’s role of blood filtration with an external dialysis machine, necessitating the creation of a permanent patent vascular access in the patient [3].

Vascular access can be achieved either through the creation of an arteriovenous fistula (AVF) or the placement of an arteriovenous graft (AVG). An AVF can be constructed by creating an anastomosis between the native vein and artery in the arm, wrist, forearm or upper arm using different surgical techniques [4]. Instead, in an AVG, the connection between the native vein and artery is achieved by means of an artificial graft, having a straight or closed-loop configuration [3]. AVGs are inevitably employed when no suitable native vein for an AVF is available [5]. Nevertheless, AVGs suffer from remarkably high failure rates (> 50% [6,7]), which are markedly associated with thrombus formation in the graft and/or progressive neointimal hyperplasia (NIH) formation leading to lumen stenosis [3,8–12]. Altered hemodynamics have been reported to play a key role in NIH initiation and progression in vascular access [13–19]. AVGs can induce hemodynamic changes at the graft-venous side, exposing the native venous wall to unphysiological flow conditions [3,15,18,20–27]. This is further supported by the clinical observation of stenosis mainly in the draining vein and at the AVG graft-venous anastomosis [16,28]. In this context, computational studies on AVGs have focused on testing different anastomotic configurations [16] and angles [29], needle positioning during hemodialysis [30] and optimising AVG designs [31,32] to minimise the occurrence of adverse hemodynamic conditions and increase graft longevity. In all the aforementioned studies, computational fluid dynamics (CFD) modelling was employed to characterise in detail the local hemodynamic environment of the AVG and test surgical and treatment scenarios *in silico*. Although hemodynamic computations rely heavily on patient-specific properties, such as vessel geometry and blood flow boundary conditions, all these studies adopted idealised models for graft and host vessel reconstruction. This is mainly due to the heterogeneity of patient-specific geometries and the limited availability and quality of imaging data, which is insufficient for tracking vessel walls at high detail over the complete length of the artery, graft and vein. Simplified boundary conditions (BCs) are also generally applied at the outlets of the distal vasculature. Flow in this region is often considered negligible and set to zero [16,30,33–35], or modelled without accounting for the peripheral vasculature response, with a flow-split condition applied between the distal vasculature and the graft [29,32,36]. Very few studies have attempted to employ representative pathophysiological conditions (i.e. lumped-parameter models), albeit on geometries where some morphological characteristics are preserved (i.e. vessel centrelines), but vessels of constant diameter are superimposed [37]. This simplification may lead to misinterpretation of the hemodynamic results, as it fails to capture the patient-specific local hemodynamic environment by overlooking vessel wall irregularities and diameter variations. Most published studies also lack validation and often focus on a limited set of near-wall hemodynamic indices — primarily low and highly oscillating wall shear stress (WSS) — while disregarding other metrics linked to vascular dysfunction, such as transverse WSS (transWSS) [38] and the topological shear variation index (TSVI) [39,40]. Furthermore, altered hemodynamics are typically not linked with regions exhibiting NIH in clinical follow-up data.

In this study, we used a retrospective dataset consisting of contrast-enhanced computed tomography angiography (CTA) scans and Doppler ultrasound (DUS) images from a patient with a brachiocephalic closed-loop AVG to develop a patient-specific modelling workflow for AVG hemodynamics. We reconstructed the patient-specific, three-dimensional (3D) geometry from CTA scans to a great level of detail – including the proximal segments of the ulnar, radial and interosseous common arteries supplying the forearm – and applied dynamic boundary conditions (e.g. three-element Windkessel model) at the arterial outlets. We then performed CFD simulations, validated the results with the patient’s clinical data and computed hemodynamic parameters such as turbulent kinetic energy, helicity descriptors, and WSS-derived indices that have been linked to AVG dysfunction. This comprehensive analysis of the hemodynamics in the AVG allowed us to identify regions presenting NIH, in agreement with the 4-month follow-up fistulogram available for this patient.

## 2. Materials and Methods

### 2.1 Clinical data

Medical imaging data from a patient with a patent left-arm brachiocephalic closed-loop AVG were acquired at Yale New Haven Hospital (New Haven, CT, USA). The study received ethical approval from the Yale Institutional Review Board (approval number 2000032101). The loop graft was created 12 years prior to data acquisition and, at the time of imaging, included indwelling stents on the proximal arterial side and in the distal venous end of the graft from prior interventions. In May 2023, DUS of the AVG was obtained with Doppler waveform recording at the left brachial artery level and the proximal edge of the arterial stent. A CTA scan of the left arm was acquired two days after the DUS images. A non-occlusive degree of stenosis was observed at the cephalic vein. A 4-month follow-up fistulogram was performed with balloon angioplasty to treat regions of occlusion at the proximal edge of the arterial stent and the cephalic vein. Images of the stenosis were only acquired and stored at the cephalic vein region. The brachial arterial systolic pressure was measured as 110 mmHg, and the central venous pressure as 8 mmHg.

### 2.2 Patient-specific geometry and meshing

The patient-specific geometry was semi-automatically segmented from the available CTA scan in ScanIP (Synopsys Inc., CA, USA) and Meshmixer (Autodesk Inc., CA, USA) using automatic thresholding and manual smoothing operations. Figure 1 shows the 3D reconstructed computational domain, including the brachial artery (inflow), the anastomosis/juxta-anastomosis region, the closed-loop AVG with two indwelling stents, and the cephalic vein region (outflow).

**Figure 1.**
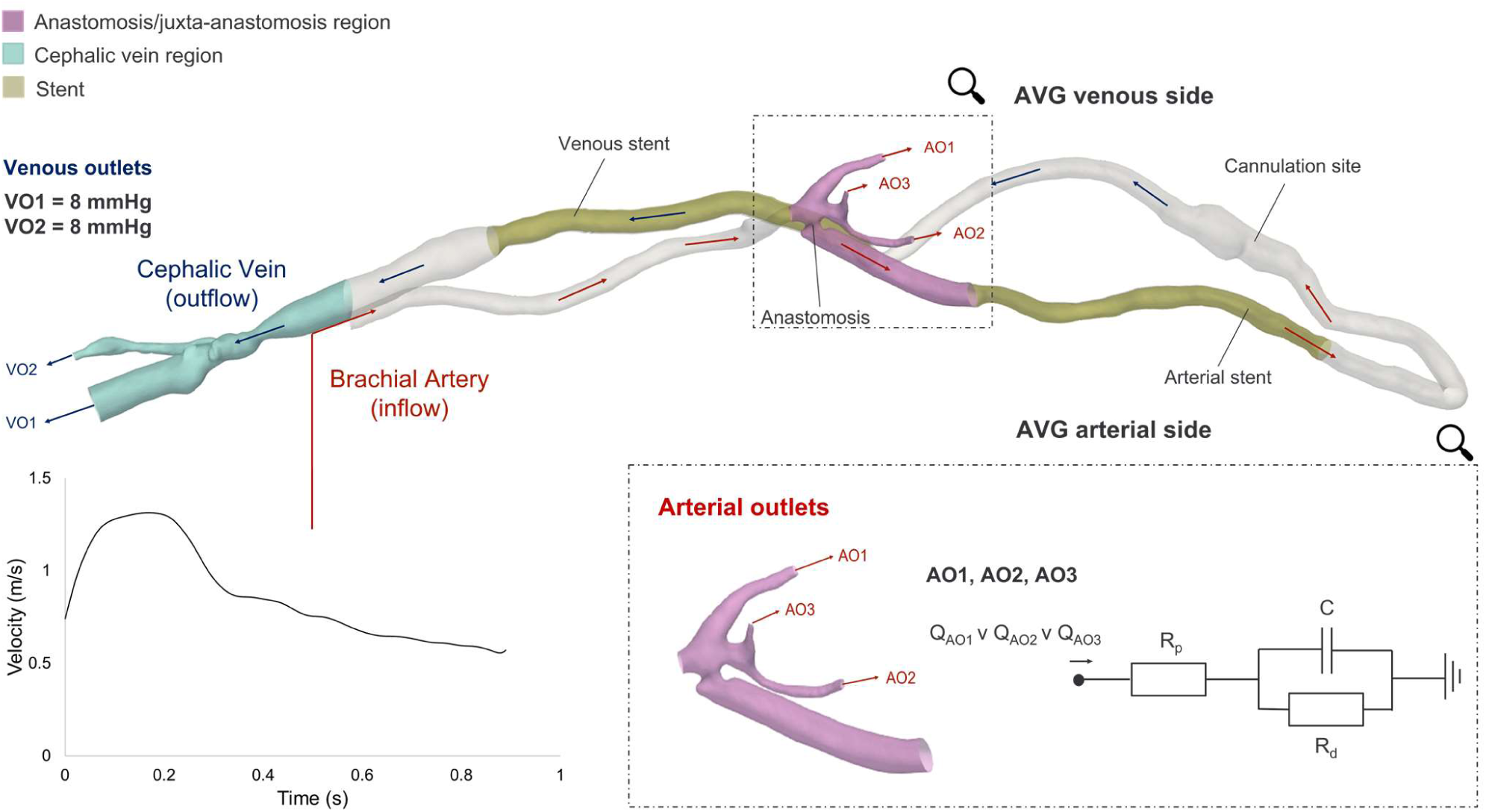
3D reconstructed patient-specific computational domain. The arrows indicate blood flow direction and distinguish between arterial (red) and venous (blue) pathways. The manually extracted patient’s DUS velocity waveform was applied as inlet boundary condition at the brachial artery. Three-element Windkessel models were implemented at the arterial outlets of the domain (AO1, AO2 and AO3), and their parameters (R_p_, R_d_ and C) were calibrated through a lumped-parameter 0D model of the computational domain. Q_AO1_, Q_AO2_ and Q_AO3_ refer to the flow rates through the respective outlets determined by their respective cross-sectional areas. A static pressure of 8 mmHg was imposed at the venous outlets (VO1 and VO2).

The vessel diameters within the indwelling stents were measured from the DUS images due to artefacts caused by the stent metallic wire mesh. The brachial artery branches into the ulnar, radial and interosseous common arteries distal to the anastomosis and their proximal segments, which supply blood to the forearm, were defined as arterial outlets in the simulations, noted as AO1, AO2 and AO3 in Figure 1. The cephalic vein also bifurcates in the upper arm, forming two venous outflow branches noted as VO1 and VO2 in Figure 1. The inlet, three arterial outlets and two venous outlets were truncated perpendicularly to the vessel centrelines.

Meshing was performed in Fluent 2023 (Ansys Inc., PA, USA) using tetrahedral elements with proximity and curvature refinement. Eight inflation layers were used with a uniform growth rate of 1.2 and a first cell height corresponding to a y+ ≤ 5. A mesh independence study was used to determine the appropriate mesh resolution using a Grid Convergence Index (GCI) approach [41–43] (*Table 1 in Supplementary Material*).

**Table 1.** Helicity descriptors (h_1_, h_2_ and h_4_) across the whole domain and ROIs.

| Helicity<br>descriptors | Whole domain |  |  | Anastomosis/juxta-anastomosis region |  |  | Arterial stent |  |  | Cephalic vein region |  |  |
| --- | --- | --- | --- | --- | --- | --- | --- | --- | --- | --- | --- | --- |
| | $h_1$ | $h_2$ | $h_4$ | $h_1$ | $h_2$ | $h_4$ | $h_1$ | $h_2$ | $h_4$ | $h_1$ | $h_2$ | $h_4$ |
|  | -5.33 | 97.34 | 0.05 | -47.21 | 251.42 | 0.19 | 83.21 | 185.51 | 0.45 | -7.56 | 40.23 | 0.18 |

### 2.3 Boundary Conditions

The applied boundary conditions are summarised in Figure 1. The velocity waveform recorded at the brachial artery level was manually extracted from the DUS images and imposed as inlet boundary condition. The resulting waveform was firstly interpolated in MATLAB (MathWorks, MA, USA) – using spline interpolation – to timesteps of 1 ms matching the simulation and applied as an inlet BC assuming a flat velocity profile. A static pressure of 8 mmHg was imposed at the venous outlets corresponding to the central venous pressure measurement.

The mean flow rate leaving the arterial outlets was determined from the available DUS velocity waveform recorded at the proximal edge of the arterial stent and the corresponding cross-sectional area measured from the CTA scan. The mean intra-AVG flow was found to be 73% of the total inflow. Therefore, the remaining 27% of the flow was directed to the arterial outlets. Three-element Windkessel (WK3) models were implemented at the arterial outlets to accurately replicate peripheral vascular response. To calibrate the WK3 parameters, a lumped-parameter 0D model of the computational domain was generated in 20-sim (Controllab, Enschede, NL).

The computational domain was divided into six sections (e.g. inlet, anastomosis, arterial outlets, arterial side graft, venous side graft and venous restriction proximal to the outflow), each represented by a single resistor and inductor in series to represent the geometric resistance (R_i_) and inertance (L_i_) of each vessel segment (Figure 1 *in Supplementary Material*). To determine R_i_ for each section, steady-state simulations of the domain across a representative flow range of the inflow waveform were performed using zero-pressure outlets at all arterial and venous outlets. At each flow rate, the pressure drop across each section was evaluated using cross-sectional planes at the proximal and distal ends of the section. The pressure drop was plotted as a function of the flow rate for each section, and a quadratic curve was fitted to each. The resulting quadratic expressions were then applied to characterise the resistor within the 0D model. Inertance values (L_i_) were derived using an expression (Equation 1) for large arteries of length l_i_ and cross-sectional area A_i_:

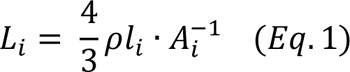

where ρ is blood density [44]. A transient CFD simulation with zero-pressure outlets was then performed using the inlet flow rate waveform computed previously. The WK3 parameters R_p_, R_d_ and C were manually tuned (R_p_ = 10, R_d_ = 15, C = 0.022) to achieve a systolic inlet pressure of 110 mmHg and a mean arterial flow output of 27% (within an error band of ±5%) of the inlet flow in the 0D model. In the absence of clinical data to prescribe a patient-specific flow split amongst the three arterial outlets, the same WK3 parameters were applied at each arterial outlet, allowing their respective cross-sectional areas to determine the flow split.

### 2.4 Computational fluid dynamics simulations

Transient simulations were performed in CFX 2023 (Ansys Inc.) using timesteps of 1 ms until cyclic periodicity was reached. This was defined as < 1% change in peak systolic pressure between subsequent cycles and was achieved after three cycles in each case with a suitable initialisation. The Reynolds-Averaged Navier-Stokes and continuity equations were solved numerically using the implicit, second-order backward-Euler method and a root-mean-square residual target of 10^-5^ for all equations within each timestep. Walls were modelled as rigid with a no-slip condition as time-resolved anatomical images were not available to model patient-specific vessel compliance.

Blood was modelled as an incompressible, non-Newtonian fluid using the Tomaiuolo formulation [45] of the Carreau-Yasuda viscosity model and a fluid density of 1056 kg/m^3^. The estimated [46,47] peak Reynolds number of 5239 exceeded the critical [46] Reynolds number of 2676, therefore the k-ω Shear Stress Transport (SST) Reynolds-averaged turbulence model was deployed using a low turbulence intensity (1%) at the inlet and all outlets [48].

### 2.5 Validation with clinical data

To validate the simulation results, the CFD-calculated volumetric flow curve obtained at the location of the proximal edge of the arterial stent was compared with the reference DUS-derived flow curve at the same location. The reference volumetric flow curve was obtained by multiplying the extracted velocity waveform from the DUS image with the cross-sectional area at the corresponding location in the AVG, derived from the patient-specific 3D reconstruction. Differences between the DUS-derived and the CFD-computed flow curves were calculated.

### 2.6 Hemodynamic analysis

The hemodynamic analysis focused on the whole domain and three regions of interest (ROIs): the anastomosis/juxta-anastomosis region, the arterial stent and the cephalic vein regions. The anastomosis/juxta-anastomosis region is commonly identified as a primary site for NIH development [49,50]. The region is also close to the arterial stent, where areas of occlusion were observed in its proximal part from the follow-up fistulogram, as well as within the cephalic vein region.

The flow field was qualitatively characterised using the velocity 3D streamlines, and velocity fluctuations were quantified to resolve the turbulent nature of the flow. Then, a comprehensive analysis of hemodynamic markers linked to AVG dysfunction was performed (turbulent kinetic energy, helicity descriptors and near-wall hemodynamics, *Table 2 in Supplementary Material*).

**Table 2.**
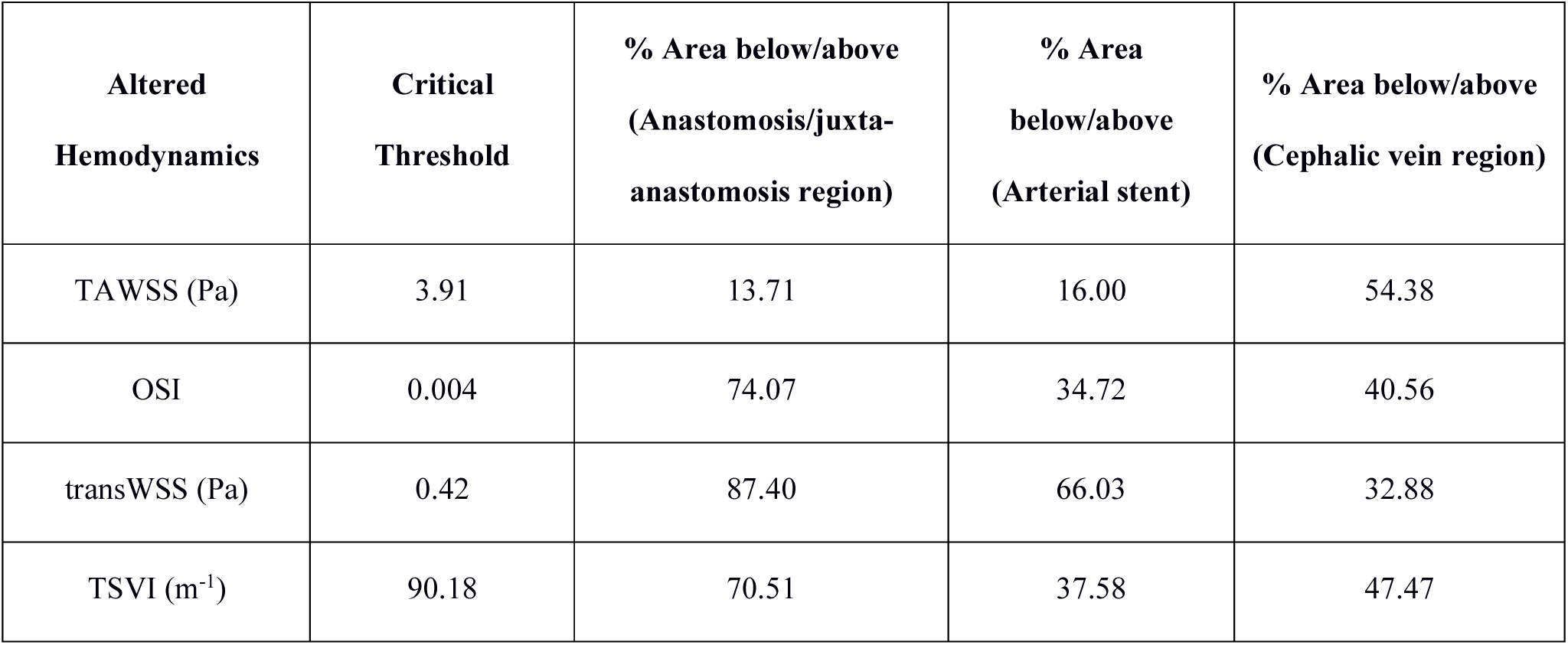
Patient-specific critical thresholds for each hemodynamic index and percentage of luminal surface in the ROIs subjected to values below or above these thresholds.

| **Altered Hemodynamics** | **Critical Threshold** | **% Area below/above (Anastomosis/juxta-anastomosis region)** | **% Area below/above (Arterial stent)** | **% Area below/above (Cephalic vein region)** |
| --- | --- | --- | --- | --- |
| TAWSS (Pa) | 3.91 | 13.71 | 16.00 | 54.38 |
| OSI | 0.004 | 74.07 | 34.72 | 40.56 |
| transWSS (Pa) | 0.42 | 87.40 | 66.03 | 32.88 |
| TSVI ($\text{m}^{-1}$) | 90.18 | 70.51 | 37.58 | 47.47 |

The turbulent kinetic energy (TKE) (*Table 2 in Supplementary Material*), whose intensity has been correlated with disease and particularly wall remodelling [49], was computed at peak systole.

Helical flow was investigated in terms of net amount and topology. Helical flow structures are recognised to suppress flow disturbances [51–56] and reduce the risk of failure in AVGs [57]. The cycle-averaged helicity (h_1_), helicity intensity (h_2_) – quantifying the net amount and the intensity of helical flow, respectively – and the unsigned (h_4_) helical rotation balance – indicating the presence of a dominant direction of rotation of helical blood structures were calculated (*Table 2 in Supplementary Material*). The local normalised helicity (LNH) [51] (*Table 2 in Supplementary Material*) was employed to visualise the helical blood flow structures in the whole computational domain.

Near-wall hemodynamics were analysed in terms of WSS-based indices accounting for the magnitude (time-averaged WSS, TAWSS), oscillatory (oscillatory shear index, OSI) and multidirectional nature (transverse WSS, transWSS) [58–65] of the WSS field over the cardiac cycle (*Table 2 in Supplementary Material*). All near-wall metrics have been reported in literature as having an impact on vascular remodelling: regions on the vessel lumen subjected to low values of TAWSS [66–68] are commonly reported as preferred sites for re-occlusion [62–64,69] due to a stimulated proatherogenic endothelial phenotype. Luminal areas subjected to high OSI values [70] coincide with disturbed flow and favour pathogenic mechanisms [62–64]. High values of transWSS (*Table 2 in Supplementary Material*) can co-locate with regions more prone to wall remodelling [60–64]. Furthermore, an analysis of the WSS topological skeleton was conducted through an Eulerian-based method relying on the divergence of the WSS vector field [71]. More specifically, the Topological Shear Variation Index (TSVI) (*Table 2 in Supplementary Material*) was used to evaluate the heterogeneity of the local contraction/expansion action exerted by the WSS on the AVG domain along the cardiac cycle [39,40]. Recent studies on the aorta, carotid bifurcations and coronary arteries reported that high values of TSVI were linked with vascular dysfunction and areas more susceptible to remodelling [72–76]. As in previous studies, the near-wall hemodynamic indices results were used to determine thresholds for altered hemodynamics (low/high values) [54,62,65,77]. To achieve this, the 33^rd^ percentile (low values) or the 66^th^ percentile (high values) of the distribution were computed, and luminal regions were considered prone to NIH development if they exhibited values lower or higher than the threshold [62]. For the whole domain and ROIs, the percentage of surface area exposed to lower values of TAWSS or higher values of OSI, transWSS and TSVI than the defined thresholds was quantified. The co-localisation between the identified “critical” (e.g. more prone to remodelling) areas and the actual remodelling observed from the available 4-month follow-up fistulogram was assessed by visual inspection.

## 3. Results

### 3.1 Validation with medical data

Figure 2 compares the flow waveform extracted from the DUS image acquired at the proximal edge of the arterial stent to the one obtained from the CFD simulation at the same location.

**Figure 2.**
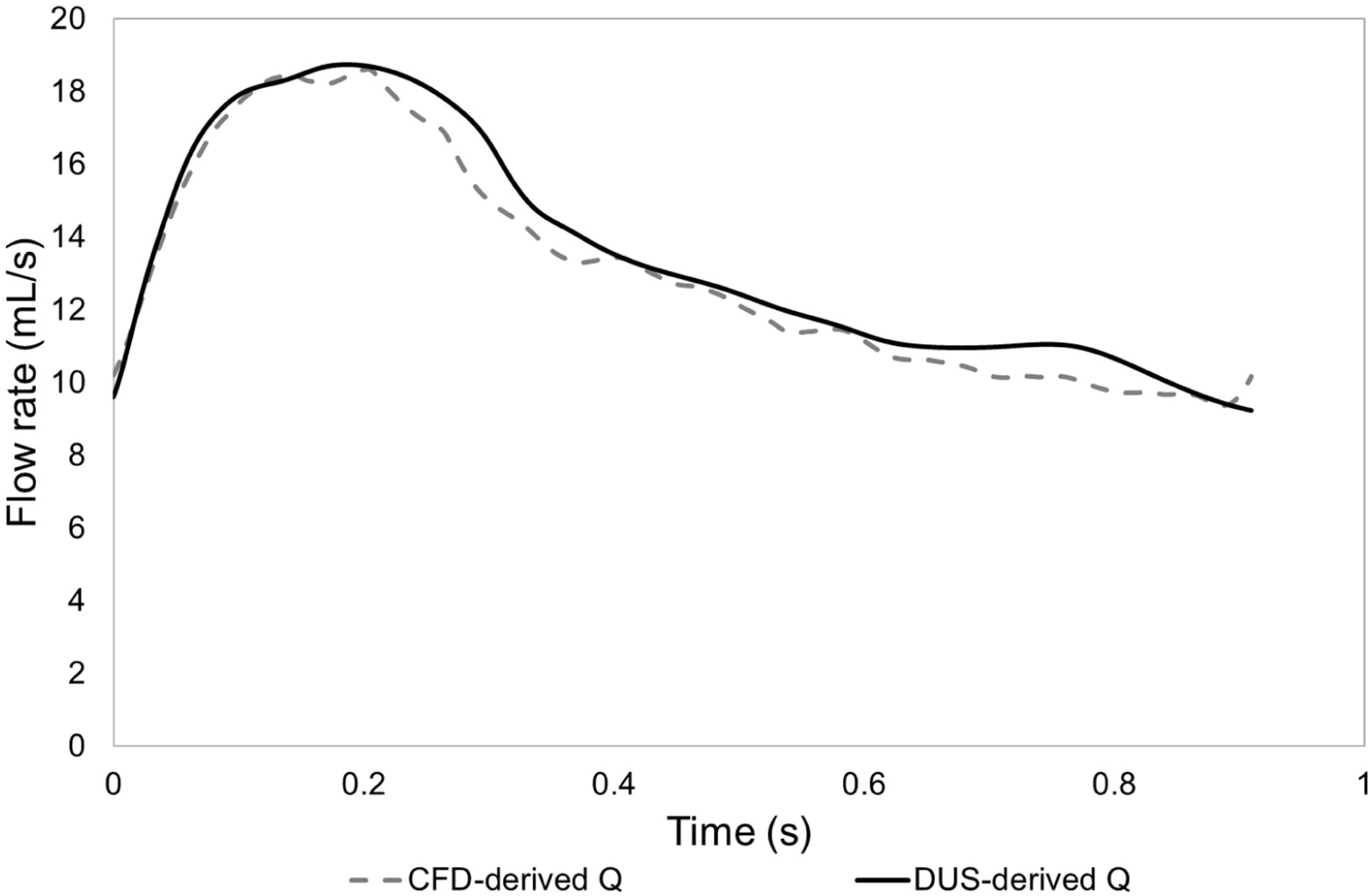
Flow waveforms derived from the DUS image (DUS-derived Q) and obtained from the CFD simulation (CFD-derived Q) at the proximal edge of the arterial stent.

It can be observed that the two flow waveforms match qualitatively (Figure 2). Quantitatively, the mean flow rate percentage difference with respect to the DUS-derived flow curve is −3.46%, with a standard deviation of 3.12%. These differences are below 5%; hence, the patient-specific geometry reconstruction and the setting of the pathophysiological boundary conditions can be considered sufficiently accurate to replicate the *in vivo* patient’s hemodynamic condition.

### 3.2 Velocity streamlines, fluctuations, TKE and NIH co-localisation

Figure 3 shows the blood flow velocity streamlines at the systolic peak. Highly disturbed flow is present throughout the whole AVG, with vortical structures forming at the anastomosis, at the cannulation site, proximally to the cephalic vein and within the cephalic vein region. Helical streamlines can be observed in the juxta-anastomosis region, while well-aligned streamlines are present in the distal part of the stent (Figure 3).

**Figure 3.**
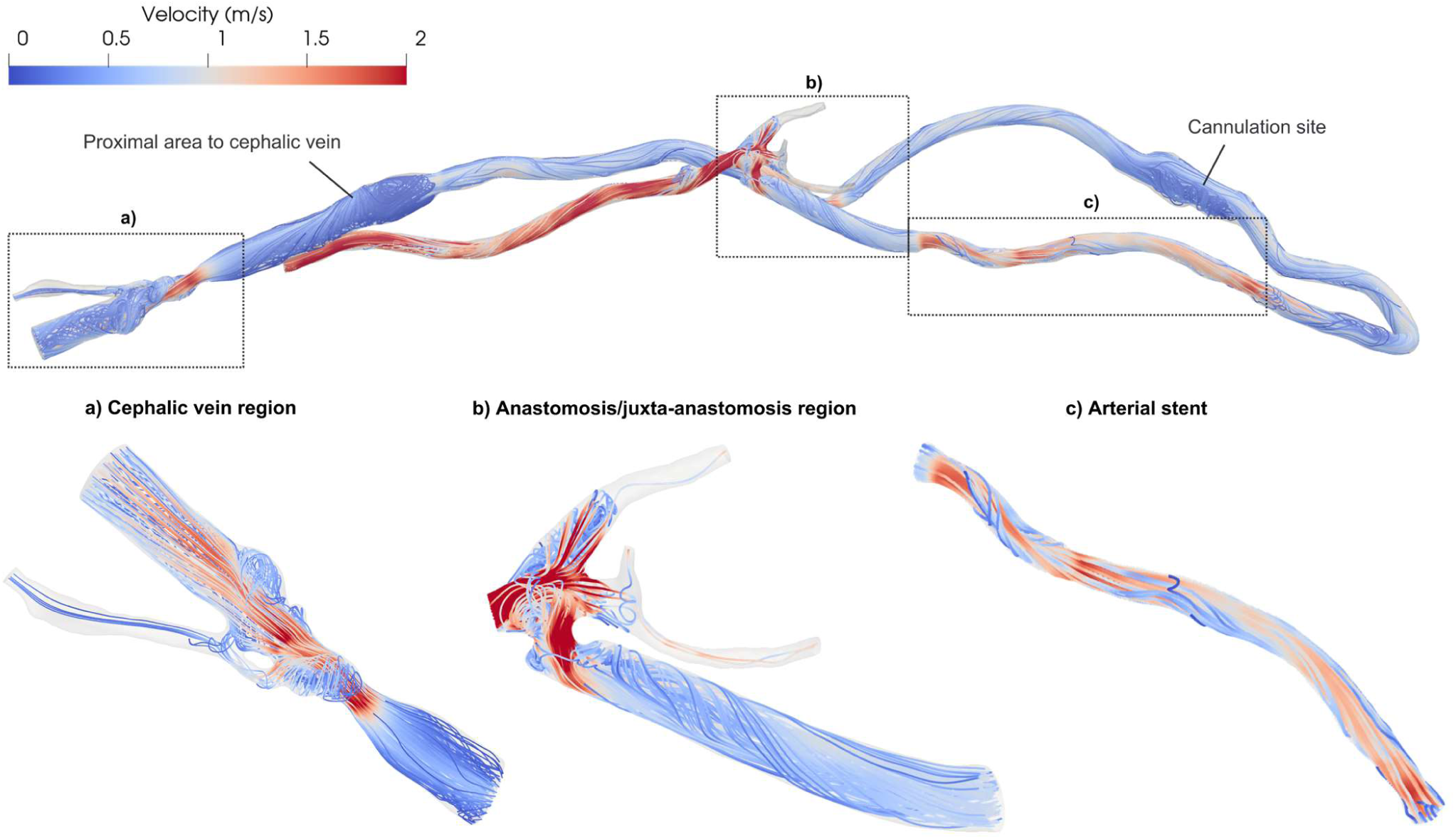
Velocity streamlines at peak systole in the whole domain and ROIs.

This highly disturbed flow is also reflected in the distributions of the streamwise velocity fluctuations (*u′*) at peak systole (Figure 4), defined as the deviations of the instantaneous values of the x-velocity component from the cycle-averaged velocity. *u′* values reach 78% of the cycle-averaged inlet velocity in the anastomosis/juxta-anastomosis region, 26% in the cephalic vein, and 31% in the arterial stent, indicating the presence of significant turbulent motion in these regions. Therefore, the turbulent kinetic energy was analysed to capture the fluctuations of all three velocity components, providing a more comprehensive picture of the turbulent flow patterns therein.

**Figure 4.**
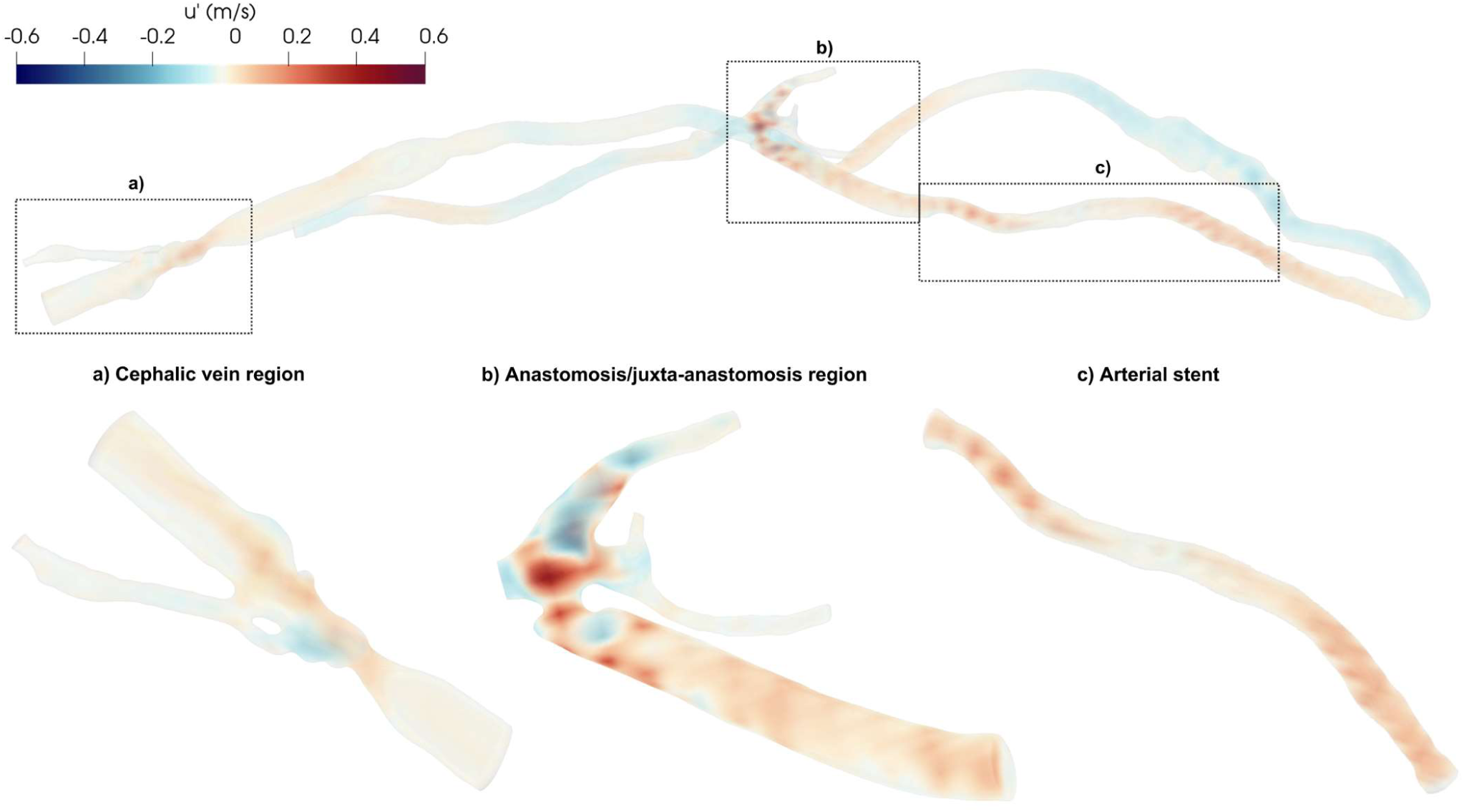
Velocity fluctuations of the x-component (*u’*) at peak systole in the whole domain and ROIs.

Figure 5 shows the volume rendering of TKE at peak systole for the whole AVG model and each ROI. The highest TKE values (up to 351.87 J/m^3^) are found at the anastomosis/juxta-anastomosis region, with lower values up to 45.43 and 61.52 J/m^3^ in the arterial stent and cephalic vein regions, respectively. Most of the remaining computational domain exhibits negligible values of TKE, reflecting the absence of significant turbulent motions (Figure 5).

**Figure 5.**
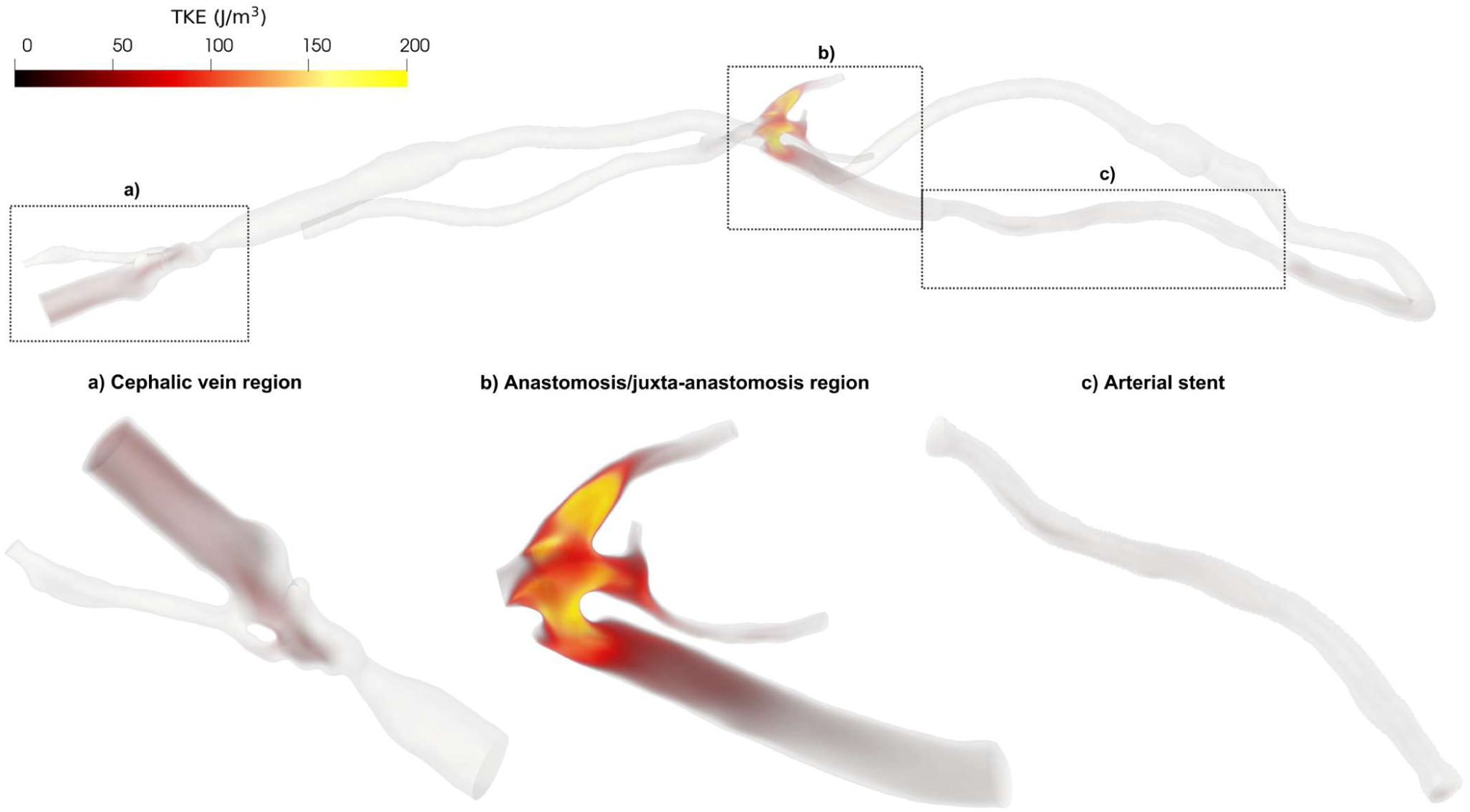
Volume rendering of TKE at peak systole for the whole domain and ROIs.

These findings are consistent with previous CFD studies on AVFs, which report highly turbulent regions (with TKE values up to 400 J/m^3^ at peak systole [49]) concentrated near the anastomosis/juxta-anastomosis location. Moreover, the occlusion reported in the 4-month follow-up fistulogram at the proximal edge of the arterial stent – corresponding to the distal part of the anastomosis/juxta-anastomosis region – might be the result of vascular wall remodelling intended to reduce flow instabilities and restore laminar flow in the AVG [50]. A similar protective mechanism might have been present at the cephalic vein region, where lower TKE values were observed at baseline alongside a non-occlusive stenosis, which later required intervention at 4-month follow-up. The hypothesis of vascular wall remodelling leading to stenosis as a protective mechanism to regularise blood flow is supported by previous studies on AVFs [50] and aligns with observations showing that vessels remodel to restore initial hemodynamic conditions and wall shear stress hemodynamics [78].

### 3.3 Helicity descriptors and NIH co-localisation

Figure 6 shows the helical blood flow patterns developing into the whole computational domain using isosurfaces of cycle-averaged LNH (at ±0.4), representing left-handed (blue) and right-handed (red) helical flow rotation [54–56]. Table 1 summarises the computed helicity-based descriptors (h_1_, h_2_, and h_4_) for the whole AVG domain and the individual ROIs.

**Figure 6.**
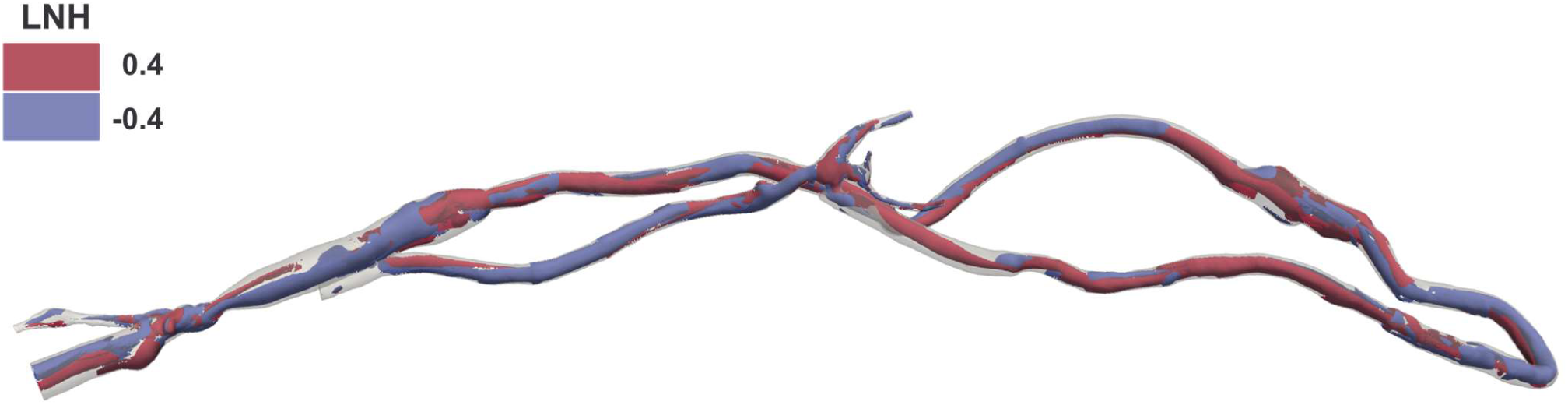
Cycle-averaged LNH in the whole computational domain. Blue and red colours indicate left-handed and right-handed helical flow rotation, respectively.

As shown in Figure 6, distinct helical flow structures are evident within the AVG domain. Quantitatively, the net amount (h_1_) – in absolute value – and the intensity of the helical flow (h_2_) are higher in the anastomosis/juxta-anastomosis and arterial stent regions than in the cephalic vein (Table 1). Although the whole domain exhibits globally balanced helical flow (h_4_ = 0.05), the arterial stent region shows a more unbalanced pattern (h_4_ = 0.45) compared to the anastomosis/juxta anastomosis and cephalic vein regions (h_4_ = 0.19 and 0.18, respectively). Unbalanced helical flow — characterised by no dominant direction of helical blood structures (right- or left-handed) — potentially leads to a more effective washout at the wall, reducing the amount of vessel wall regions exposed to low and oscillatory WSS [32]. This could explain the development of stenosis in regions with more balanced helical flow patterns, such as the cephalic vein region and the proximal edge of the arterial stent, located distally to the anastomosis region.

### 3.4 Near-wall hemodynamics and NIH co-localisation

The computed distributions of near-wall hemodynamic indices enabled the identification of patient-specific critical thresholds, as well as the percentage of surface area exposed to values above or below these thresholds in the ROIs (Table 2). Figure 7 shows the identified critical luminal areas for each hemodynamic index.

**Figure 7.**
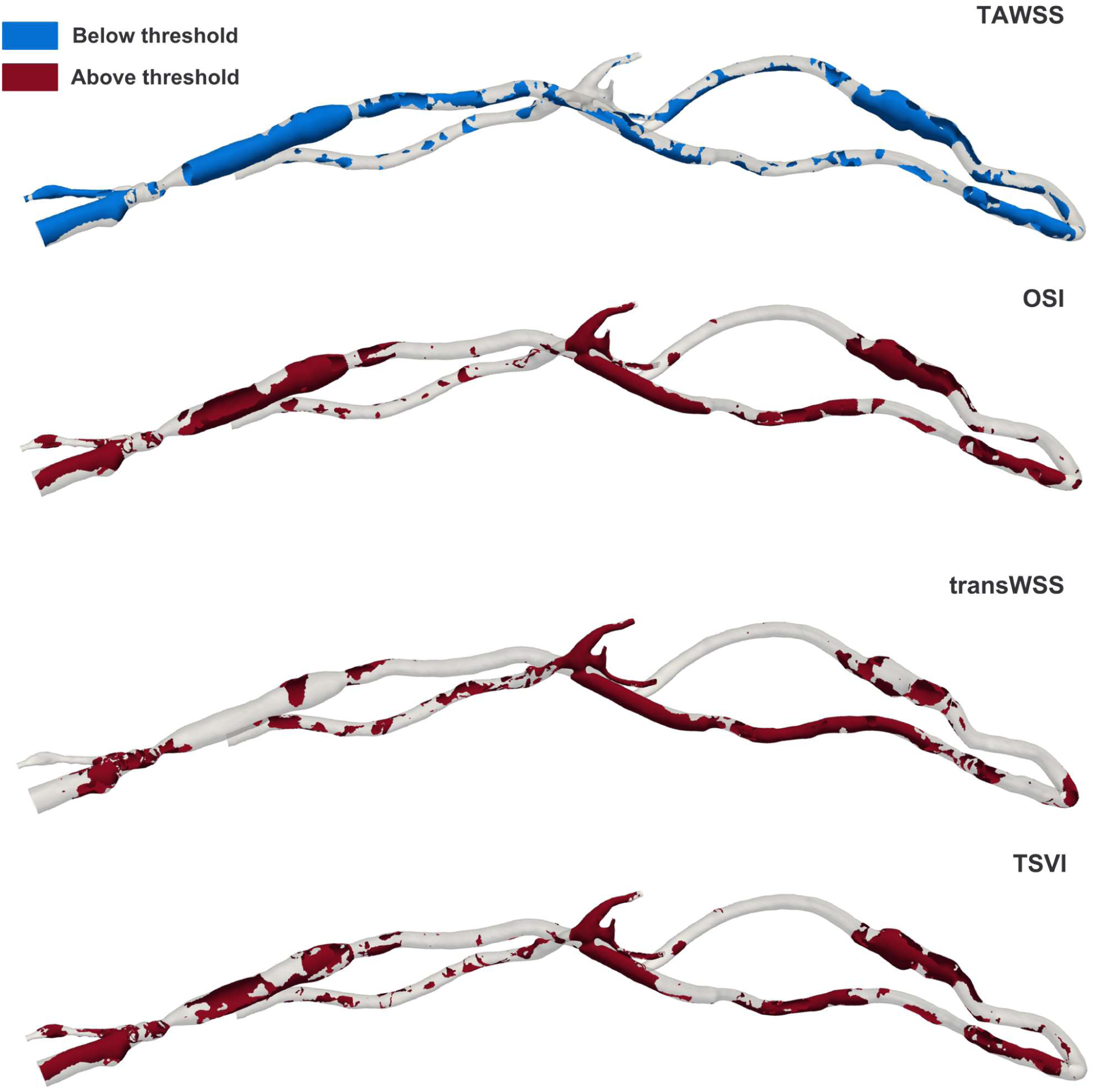
Identified critical areas – more prone to occlusion – below/above patient-specific thresholds (Table 2).

A remarkably high percentage of surface area (> 70%) is subjected to high values of OSI, transWSS and TSVI in the anastomosis/juxta-anastomosis region. In the arterial stent and cephalic vein regions, the percentages are all above 30%, reaching 66.03% for the transWSS index in the arterial stent region (Table 2). The TAWSS index depicts a relatively small percentage of critical areas in the anastomosis/juxta-anastomosis and arterial stent regions (13.71% and 16.00%, respectively), but higher than 50% in the cephalic vein region. Since regions that occluded were observed from the clinical data at the proximal edge of the arterial stent and at the cephalic vein, the transWSS index detects the highest percentage of critical areas in the arterial stent region and best co-localises qualitatively critical areas with the occlusion at the cephalic vein region (Figure 8). Moreover, compared to TAWSS, OSI and TSVI, transWSS also minimises the percentage of detected critical “false” areas at the cannulation site (Figure 1) and at the venous side of the AVG proximal to the cephalic vein region (Figure 7). Nevertheless, extensive critical areas in the whole anastomosis/juxta-anastomosis region (Figure 7) are wrongly depicted by OSI, transWSS and TSVI due to that area being highly turbulent (Figure 5).

**Figure 8.**
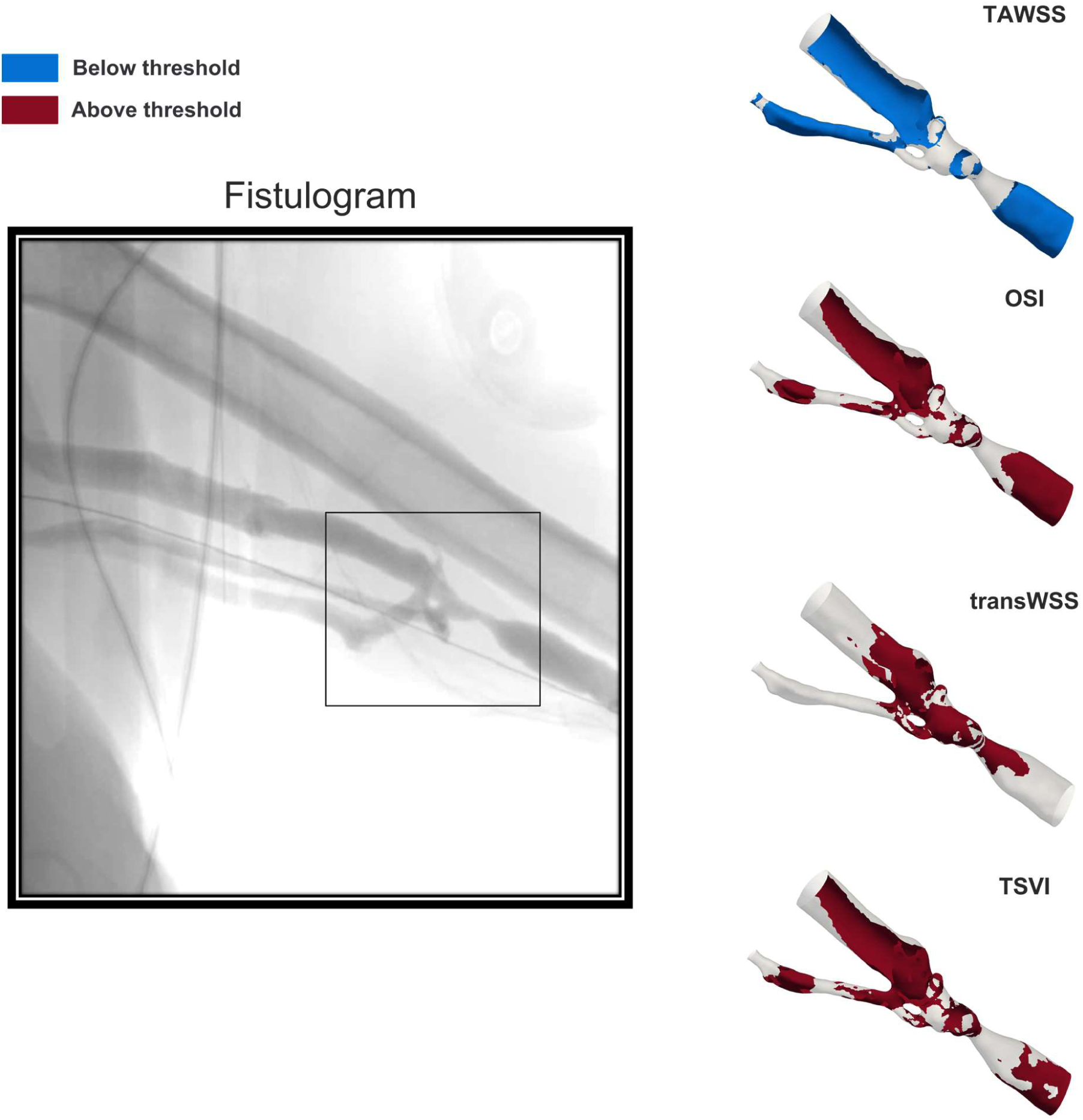
A 4-month follow-up fistulogram image of the cephalic vein region identified critical areas at the same location by the different near-wall hemodynamic indices. The transWSS index best co-localised qualitatively with the observed occlusion.

This work was motivated by the need for validated, patient-specific CFD simulations of AVGs to enable the translation of hemodynamic markers into clinical biomarkers of NIH progression. Most computational studies on AVGs rely on idealised models, with simplified BCs applied at the arterial outlets, and fail to thoroughly investigate the co-localisation of altered hemodynamics with post-operative NIH development.

To address this, we leveraged a retrospective dataset of contrast-enhanced CTA scans and DUS images to perform what is, to the best of the authors’ knowledge, the first analysis on a fully patient-specific closed-loop AVG geometry. We also implemented realistic, pathophysiological boundary conditions (i.e. lumped-parameter conditions) at the arterial outlets of the domain to account for the response of the peripheral vasculature. The detailed patient-specific geometry reconstruction and lumped-parameter BCs applied at the arterial outlets successfully captured the patient’s *in vivo* hemodynamic conditions. A comprehensive analysis of the CFD results – including turbulent kinetic energy, helicity and near-wall hemodynamic descriptors (TAWSS, OSI, transWSS and TSVI) – allowed us to investigate whether altered hemodynamics in the AVG co-localised with regions exhibiting NIH growth, as observed from a 4-month follow-up fistulogram.

Highly turbulent regions at baseline were found to coincide with regions presenting NIH at 4-month follow-up. The anastomosis/juxta-anastomosis region was identified as the most turbulent region in the whole domain, and occlusion appeared in its distal part, in correspondence to the proximal portion of the arterial stent. The cephalic vein region – undergoing occlusion at 4-month follow-up and presenting non-occlusive stenosis at baseline – exhibited lower values of TKE potentially due to a protective remodelling process that tends to progressively regularise the blood flow. Inward vessel remodelling might have been regions, since previous experimental studies have demonstrated that oscillating flow adjacent to the vessel wall induces a proliferative, pro-inflammatory and pro-oxidant state for the endothelial cells, resulting in impaired vascular tone regulation [79,80]. More balanced helical blood flow structures, exhibiting a dominant direction (right- or left-handed), were observed at baseline in regions that later developed NIH at the 4-month follow-up. These structures may contribute to a less effective wall washout and an increase in areas exposed to near-wall disturbed flow indices [32]. A significant finding of this study is that areas subjected to high transWSS best co-localised qualitatively with the occlusion at the cephalic vein region compared to other commonly studied near-wall hemodynamic indices. Most studies on AVGs to date have limited their analysis to TAWSS and OSI [32,37,49,50,81], disregarding other potential important predictors of NIH growth. The transWSS index detected the highest percentage of critical areas in the arterial stent region – where occlusion is present at its proximal part – and minimised the identification of critical “false” areas elsewhere. Even though extensive critical areas in the anastomosis/juxta-anastomosis region were incorrectly depicted by transWSS, it is important to note that this region is reasonably close to the proximal portion of the arterial stent.

For completeness, additional simulations with different outlet boundary conditions were conducted to assess the impact of simplified conditions at the arterial outlets, such as prescribed flow-split or zero-flow conditions. We found that these may have a limited impact on certain predicted hemodynamic metrics, including TKE and helicity descriptors (*see Tables 3 and 4 in Supplementary Material*). However, statistically significant differences in near-wall hemodynamic indices were observed (Figure 2 *in Supplementary Material*). Notably, the flow-split condition consistently identified the same critical regions across hemodynamic indices (> 90% Similarity Index, *Equation 1 and Table 5 in Supplementary Material*), aligning with findings from previous AVG studies where critical regions were identifiable despite some statistical differences in WSS-related indices [65]. This consistency suggests that the flow-split condition may serve as a practical boundary condition strategy for specific applications, offering the advantage of avoiding time-intensive tuning of 0D models. However, the lumped-parameter conditions used in this study provide an essential capability that flow-split conditions lack: the flexibility to account for scenarios involving retrograde flow to and from the forearm. This adaptability makes the lumped-parameter approach used here particularly robust, suitable for a wider range of hemodynamic conditions, and reflective of patient-specific dynamics.

It should be noted that our dataset is limited to one patient; hence, analyses on a larger cohort would strengthen the results and discussion. Moreover, the simulated AVG model does not represent the patient’s condition immediately after intervention. A prospective cohort of patients with data acquired immediately after intervention and at subsequent defined time points would enable a more detailed investigation of the evolution of the hemodynamic results and their link with NIH progression. All simulations assumed rigid walls, as no medical data were available to model patient-specific vessel compliance. The impact of this assumption on the hemodynamic results and the co-localisation with regions presenting NIH growth should be investigated in future work. Lastly, the validation of the CFD results was performed using DUS measurements. These measurements are one-dimensional and refer to an isolated region of the vessel. DUS accuracy also depends on the intrinsic properties of the ultrasound beam and the measurements also suffer from operator dependence. This source of error is challenging to minimise, and even if possible, it would be reasonable to expect the same conclusion in terms of mean flow rate percentage difference with the medical data. However, to the best of our knowledge, this is the first study to attempt validation of CFD results on AVGs with patient-specific clinical data.

This study employed a retrospective dataset of CTA scans and DUS images to develop a novel, patient-specific modelling workflow for a closed-loop AVG. The available CTA scans allowed an accurate reconstruction of the AVG to a level of detail that has not been reported previously in the literature. Additionally, pathophysiological representative boundary conditions (i.e. lumped-parameter conditions) – normally simplified in other studies – were applied at the arterial outlets to model the flow to and from the distal vasculature. Patient-specific CFD simulations were then performed, validated with the patient’s clinical data and used to co-localise the altered hemodynamics in terms of turbulent kinetic energy, helicity descriptors and near-wall hemodynamic indices (TAWSS, OSI, transWSS and TSVI) with regions presenting NIH growth observed from a 4-month follow-up fistulogram.

The results reveal that highly turbulent areas and the presence of balanced helical flow structures co-localise with areas of NIH growth. Moreover, among the studied near-wall hemodynamic indices linked to vascular dysfunction (i.e. TAWSS, OSI and TSVI), transWSS seems a potentially stronger predictive marker of NIH development. Areas subjected to high transWSS greatly co-localise with areas undergoing inward remodelling at the cephalic vein region, and a high percentage of critical areas is depicted in the stented region where occlusion occurs in the proximal part. Although this study only considers a single patient, it proposes a computational workflow based on CTA and DUS imaging data that can be applied to a larger cohort, paving the way for a detailed investigation of the clinical biomarkers of NIH in vascular accesses.

## Statements and Declarations

## Data Availability

All data produced in the present study are available upon reasonable request to the authors.

