## Supplementary Material for "Patient-specific hemodynamic markers co-localise with regions of neointimal hyperplasia in an arteriovenous graft"

#### Tables

Three successively refined meshes were used to perform rigid-wall transient simulations with consistent boundary conditions (see Subsection 2.3 in the Manuscript). Mesh element count approximately doubled between successive refinements. Simulations were initialised using a previously converged simulation, and three further cycles were run. Less than a 1% difference in systolic and diastolic pressures was observed between these three cycles for each mesh. The same settings described in the main manuscript (see Subsection 2.4) were adopted for the different simulations.

As shown in Table 1, differences between medium and fine meshes did not exceed 1.93% in absolute value for all the metrics, whereas differences up to almost 25% were observed between the coarse and medium meshes.

Results using the Grid Convergence Index (GCI) approach [1–3] confirmed and strengthened these findings. The GCI was calculated as a percentage using the following equations, where c, m and f correspond to quantities from the coarse, medium and fine meshes, respectively:

$$r_{f,m} = \left(\frac{N_m}{N_f}\right)^{\frac{1}{3}} \approx r_{m,c} = \left(\frac{N_c}{N_m}\right)^{\frac{1}{3}}$$

$$r = \frac{r_{f,m} + r_{m,c}}{2}$$

$$p = \frac{\ln\left(\frac{|f_c - f_m|}{|f_m - f_f|}\right)}{\ln(r)}$$

$$E_{f,m} = \frac{\left(\frac{|f_m - f_f|}{f_f}\right)}{r^p - 1} \qquad E_{m,c} = \frac{\left(\frac{|f_c - f_m|}{f_m}\right)}{r^p - 1}$$

$$GCI_{f,m} = F_s \left|E_{f,m}\right| \qquad GCI_{m,c} = F_s \left|E_{m,c}\right|$$

*N* is the number of elements in the mesh, *f* is the examined variable of interest and $F_s$ is a safety factor of 1.25 [1–3]. The GCI never exceeded 3.87% for any quantity in any mesh. However, between the medium and fine meshes, the GCI never exceeded 1.69%. For these reasons, the medium mesh was used for all further analysis in the study.

**Table 1**. Table of key metrics of interest from coarse, medium and fine meshes. Percentage difference between medium/coarse ($\%_{m,c}$) and fine/medium ($\%_{f,m}$) meshes and GCI values for the medium/coarse ($\%GCI_{m,c}$) and fine/medium meshes ($\%GCI_{f,m}$) .

| Metric | Fine | Medium | Coarse | $\%_{f,m}$ | $\%_{m,c}$ | $\%GCI_{f,m}$ | $\%GCI_{m,c}$ |
|---|---|---|---|---|---|---|---|
| Node count | 1,780,121 | 891,182 | 395,186 | 49.94 | 55.66 | / | / |
| Element count | 5,756,247 | 2,595,857 | 984,828 | 54.90 | 62.06 | / | / |
| Inflation layers | 8 | 8 | 8 | / | / | / | / |
| Refinement ratio (r) | | 0.75 | | / | / | / | / |
| Max velocity magnitude @ peak systole (m/s) | 4.33 | 4.31 | 4.27 | **0.51** | 0.93 | **0.79** | 1.43 |
| Mean velocity magnitude @ peak systole (m/s) | 0.85 | 0.85 | 0.81 | **0.01** | 4.05 | **0.00** | 0.01 |
| $WSS_{max}$ @ peak systole (Pa) | 188.00 | 191.63 | 201.00 | **-1.93** | -4.89 | **1.53** | 3.87 |
| $WSS_{avg}$ @ peak systole (Pa) | 23.03 | 22.88 | 17.11 | **0.66** | 25.21 | **0.02** | 0.85 |

| | | | | | | | |
|---|---|---|---|---|---|---|---|
| Max TAWSS (Pa) | 108.57 | 107.22 | 107.33 | **1.24** | -0.10 | **1.69** | 0.14 |
| Mean TAWSS (Pa) | 13.32 | 13.17 | 11.73 | **1.10** | 10.94 | **0.16** | 1.55 |
| Max OSI | 0.49 | 0.50 | 0.48 | **-0.69** | 3.64 | **0.20** | 1.05 |

**Table 2**. Formulas and explanations of the turbulent kinetic energy, helicity and near-wall hemodynamic descriptors investigated in the present study.

| | | |
|---|---|---|
| **Turbulence descriptors** | | |
| Turbulent Kinetic Energy (TKE) | It represents the turbulence intensity in the fluid. ρ is blood density, and u', v', w' are the fluctuating components of blood velocity, obtained by subtracting the corresponding phase-averaged velocity components from the instantaneous ones. | $TKE = \frac{1}{2}\rho((u')^2 + (v')^2 + (w')^2)$ |
| **Helicity descriptors** | | |
| Average Helicity ($h_1$) | Time-averaged value of helicity (equal to 0 in the presence of reflectional symmetry in the fluid domain) | $h_1 = \frac{1}{TV}\iint_{T\,V} \mathbf{v} \cdot \boldsymbol{\omega}\; dVdt$ |
| Average Helicity Intensity ($h_2$) | Helicity intensity, an indicator of the total amount of helical flow in the fluid domain, irrespective of direction | $h_2 = \frac{1}{TV}\iint_{T\,V} \lvert\mathbf{v} \cdot \boldsymbol{\omega}\rvert\; dVdt$ |
| Unsigned balance of counter-rotating helical flow structures ($h_4$) | Indicating the presence of a dominant direction of helical blood structures. It ranges from 0 (totally balanced) to 1 (totally unbalanced) | $h_4 = \frac{\lvert h_1\rvert}{h_2} \quad 0 \le h_4 \le 1$ |
| Local Normalised Helicity (LNH) | Normalised internal product between local velocity and vorticity vectors. The dimensionless quantity of LNH measures the (mis)alignment of the local velocity vector with respect to the vorticity vector. If positive, the fluid structures rotate along the left-handed direction; if negative, along the right-handed one. | $LNH = \frac{\mathbf{v} \cdot \boldsymbol{\omega}}{\lvert\mathbf{v} \cdot \boldsymbol{\omega}\rvert} = \cos(\gamma)$ |
| **Near-wall hemodynamic descriptors** | | |

| | | |
|---|---|---|
| Time-averaged WSS (TAWSS) | Cardiac cycle-averaged WSS vector magnitude | $\mathrm{TAWSS} = \frac{1}{\mathrm{T}}\int_0^{\mathrm{T}} \lvert\mathbf{WSS}\rvert\,\mathrm{dt}$ |
| Oscillatory Shear Index (OSI) | A measure the directional change of WSS during the cardiac cycle, accounting for the degree of flow reversal. It ranges between 0 (totally unidirectional WSS vector) and 0.5 (purely oscillatory WSS with a net magnitude of zero) | $\mathrm{OSI} = 0.5\left[1-\left(\frac{\left\lvert\int_0^{\mathrm{T}}\mathbf{WSS}\,\mathrm{dt}\right\rvert}{\int_0^{\mathrm{T}}\lvert\mathbf{WSS}\rvert\,\mathrm{dt}}\right)\right]$ |
| Transverse WSS (TransWSS) | Average of the WSS vector component acting orthogonal to the cardiac cycle-averaged WSS vector direction | $\mathrm{transWSS} = \frac{1}{\mathrm{T}}\int_0^{\mathrm{T}}\left\lvert \boldsymbol{WSS} \cdot \left(\boldsymbol{n} \times \frac{\int_0^{\mathrm{T}}\boldsymbol{WSS}\,\mathrm{dt}}{\left\lvert\int_0^{\mathrm{T}}\boldsymbol{WSS}\,\mathrm{dt}\right\rvert}\right)\right\rvert \mathrm{dt}$ |
| Topological Shear Variation Index (TSVI) | The root mean square deviation of the divergence of the normalized WSS vector with respect to its average over the cardiac cycle. It quantifies the variability of WSS contraction/expansion action exerted at the endothelium along the cardiac cycle | $\mathrm{TSVI} = \left\{\frac{1}{\mathrm{T}}\int_0^{\mathrm{T}}\left[\nabla \cdot (\boldsymbol{WSS}_{\mathrm{u}}) - \overline{\nabla \cdot (\boldsymbol{WSS}_{\mathrm{u}})}\right]^2 \mathrm{dt}\right\}^{1/2}$ |

**Table 3**. Ranges of TKE values at peak systole for the whole domain and ROIs across each BC scenario.

| TKE (J/m$^3$) | Whole domain | Anastomosis/juxta-anastomosis region | Arterial stent | Cephalic vein region |
|---|---|---|---|---|
| **WK3** | 0 – 351.87 | 0 – 351.87 | 0 – 45.43 | 0 – 61.52 |
| **Flow-split** | 0 – 454.56 | 0 – 454.56 | 0 – 60.47 | 0 – 89.48 |
| **Zero-flow** | 0 – 186.99 | 0 – 186.99 | 0 – 56.55 | 0 – 90.01 |

**Table 4**. Helicity descriptors ($h_1$, $h_2$ and $h_3$) across the whole domain and within ROIs under the different BC scenarios.

| Helicity descriptors | Whole domain | Anastomosis/juxta-anastomosis region | Arterial stent | Cephalic vein region |
|---|---|---|---|---|

| | $h_1$ | $h_2$ | $h_4$ | $h_1$ | $h_2$ | $h_4$ | $h_1$ | $h_2$ | $h_4$ | $h_1$ | $h_2$ | $h_4$ |
|---|---|---|---|---|---|---|---|---|---|---|---|---|
| **WK3** | -5.33 | 97.34 | 0.05 | -47.21 | 251.42 | 0.19 | 83.21 | 185.51 | 0.45 | -7.56 | 40.23 | 0.18 |
| **Flow split** | -5.12 | 97.90 | 0.05 | -41.50 | 249.94 | 0.17 | 77.25 | 184.77 | 0.42 | -7.44 | 41.42 | 0.18 |
| **Zero-flow** | -9.90 | 82.46 | 0.12 | -108.18 | 236.28 | 0.46 | 4.51 | 153.42 | 0.03 | -7.66 | 41.39 | 0.19 |

**Table 5**. Similarity indices to the three-element Windkessel model (WK3) case in terms of identified critical luminal areas for each hemodynamic index and BC scenario.

| **Similarity Index** | **WK3 – Flow-Split** | **WK3 – Zero-Flow** |
|---|---|---|
| TAWSS | 0.99 | 0.84 |
| OSI | 0.94 | 0.85 |
| transWSS | 0.93 | 0.76 |
| TSVI | 0.94 | 0.90 |

### Equations

The co-localisation of the identified low/high hemodynamic values on the luminal wall obtained with different BC strategies to the three-element Windkessel (WK3) one was assessed by applying the Jaccard similarity index (SI) [4,5]:

$$SI = \frac{2(Indexperc_{WK3} \cap Indexperc_{BC})}{(Indexperc_{WK3} \cup Indexperc_{BC})} \quad (Eq. 1)$$

where $Indexperc_{WK3}$ is the luminal surface area exposed to low (33$^{rd}$ percentile) or high (66$^{th}$ percentile) values when the WK3 strategy is applied at the arterial outlets of the domain and $Indexperc_{BC}$ is the luminal surface area exposed to either low or high values when other strategies are used. The SI ranges from 0 (no co-localisation) to 1 (perfect co-localisation). This metric determined whether the same critical regions observed when applying WK3 conditions at the arterial outlets could be depicted when adopting flow-split or zero-flow conditions.

### Figures

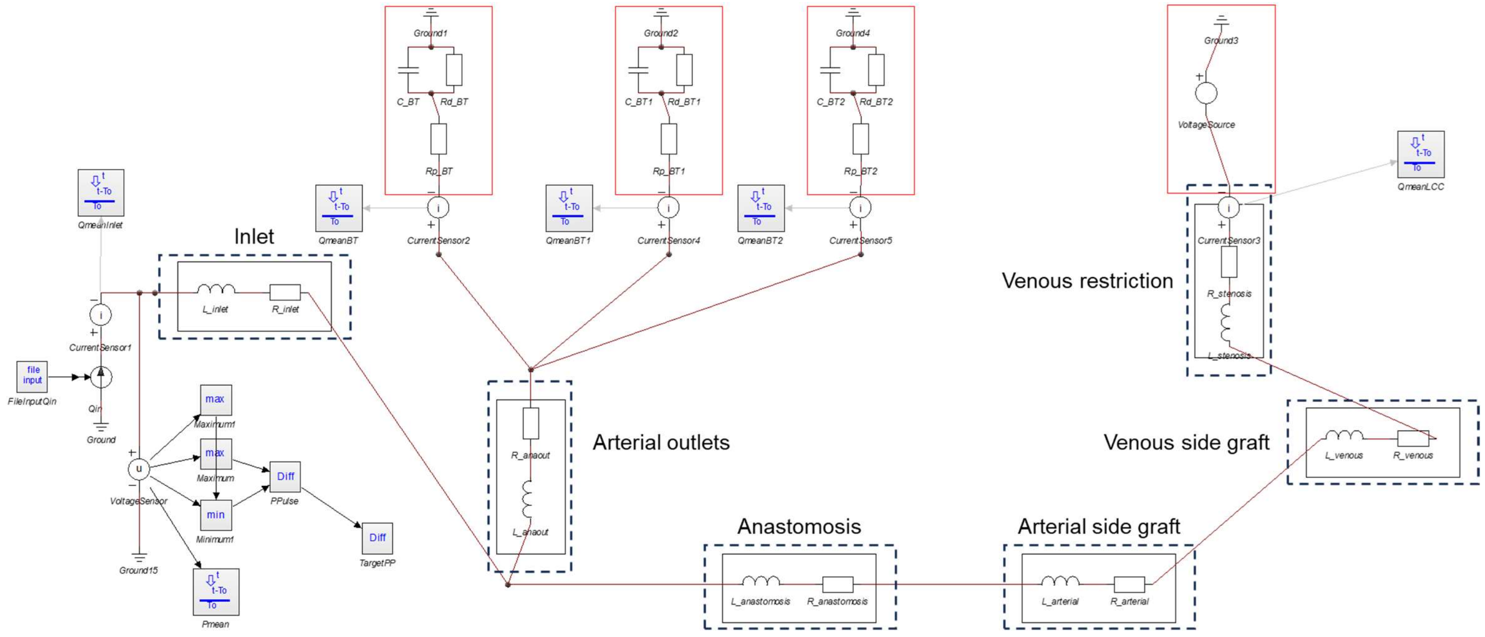


**Figure 1**. Lumped-parameter 0D model of the computational domain divided into six sections (e.g. inlet, anastomosis, arterial outlets, arterial side graft, venous side graft and venous restriction proximal to the outflow), each represented by a single resistor and inductor in series to represent the geometric resistance and inertance of each vessel segment.

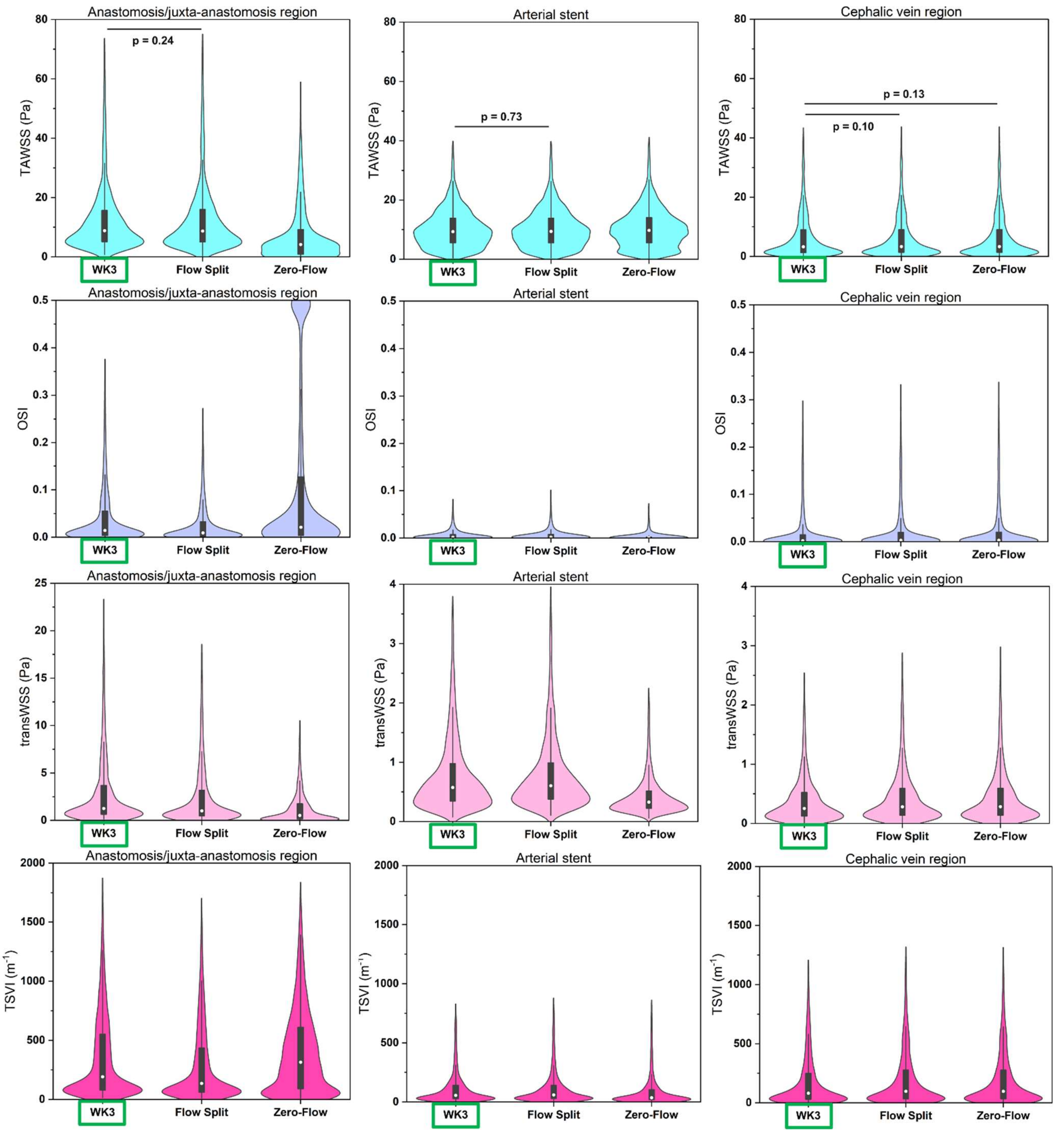


**Figure 2**. Violin plots of TAWSS, OSI, transWSS and TSVI distributions for each ROI and BC scenario. The distributions obtained under the three-element Windkessel (WK3) scenario are highlighted by a rectangle. When compared to the WK3 distribution, the majority of distributions resulted statistically significantly different ($p < 0.01$), except for the TAWSS distributions obtained in all ROIs under the flow split BC scenario and in the cephalic vein region under the zero-flow scenario.
